# DNA methylation signatures of C-reactive protein associations with structural neuroimaging measures and major depressive disorder

**DOI:** 10.1101/2020.08.12.20173567

**Authors:** Claire Green, Xueyi Shen, Anna J. Stevenson, Eleanor L.S. Conole, Mathew A. Harris, Miruna C. Barbu, Emma L. Hawkins, Mark J. Adams, Stephen M. Lawrie, Kathryn L. Evans, Rosie M. Walker, Stewart W. Morris, David J. Porteous, Joanna M. Wardlaw, J Douglas Steele, Gordon D. Waiter, Anca-Larisa Sandu, Archie Campbell, Riccardo E. Marioni, Simon R. Cox, Jonathan Cavanagh, Andrew M. McIntosh, Heather C. Whalley

## Abstract

**Background:** Inflammatory processes are implicated in the aetiology of Major Depressive Disorder (MDD); however, the relationship between peripheral inflammation, brain structure and depression remains unclear. This study investigates associations between depression, structural neuroimaging measures and two measures of inflammation: serum-based C-reactive protein (CRP) and a methylation-based score for CRP (DNAm CRP) in a large community-based sample.

**Methods:** Serum CRP and DNAm CRP were assessed for participants in Generation Scotland (N_MDD cases_ = 240, N_controls_ = 558) alongside structural brain scans (T1 and diffusion MRI). Linear regression was used to investigate associations between (i) both CRP measures and depressive symptoms, (ii) both CRP measures and structural imaging variables and (iii) inflammation x MDD interaction effects (for both CRP measures) with imaging measures.

**Results:** Increased serum CRP was significantly associated with overall MDD symptom severity and specifically with somatic symptoms-general interest (β= 0.145, P_FDR_ = 6×10^-4^) and energy levels (β= 0.101, P_FDR_ = 0.027) and also reduced entorhinal cortex thickness (β= – 0.095, P_FDR_ = 0.037). DNAm CRP was significantly associated with reduced global grey matter/cortical volume and reduced integrity of 16 white matter tracts and showed larger effect sizes (β_average_ = −0.15) compared to serum CRP across all measures (β_average_ = 0.01).

**Conclusions:** Acute measures of CRP were related to current depression symptoms, specifically somatic symptoms, whereas methylation signatures of inflammation demonstrated greater differences in global and regional brain structure. This study highlights the utility of combining serological and methylation markers to study chronic inflammation effects on the brain in psychiatric disorders.

## 1. Introduction

Major depressive disorder (MDD) is the most common mental health condition in the general population ^1^. It is a heritable disorder linked to a diminished functioning and quality of life, medical morbidity, and mortality ^2,3^ Activation of the peripheral immune system has been consistently associated with MDD and is implicated in its pathogenesis ^4-8^. Evidence has shown that markers of inflammation are upregulated in peripheral and central nervous system tissues of individuals with depression, including increased concentrations of proinflammatory cytokines and immune mediators in the cerebrospinal fluid of those with MDD compared to healthy controls ^9-12^ Furthermore, a meta-analysis of randomised control trials (n=2,370) of pro-inflammatory cytokine inhibitors showed that these treatments significantly improved depressive symptoms compared with placebo, indicating a potentially casual role of inflammation in MDD ^13^. However, despite the clear association between inflammation and mood disorder, the effects of peripheral inflammatory markers on brain structure in depression remain largely unknown.

One of the most common ways to determine peripheral inflammation is by measuring serum levels of C-reactive protein (CRP). CRP plays a key role in human inflammation and can provide a proxy estimate for inflammatory activity ^14^ It is widely used in clinical practice as a marker of inflammation and is a candidate biomarker for investigating inflammatory processes in MDD ^15^. Meta-analyses have shown that serum CRP levels are reliably elevated in MDD ^16^ and have been found to predict future development of depression as well as resistance to standard antidepressant therapies ^6,17,18^

Despite evidence that peripheral inflammation is related to MDD, and that CRP can disrupt the BBB and activate local inflammatory pathways, there is little evidence of associations between elevated serum CRP in MDD and alterations in brain structure^19-21^. One exception is a large imaging study by Opel and colleagues that did report significantly increased CRP levels in association with reduced grey matter volume in 514 patients with MDD compared to 359 healthy controls^22^.

Although serum CRP is viewed as a proxy measure of inflammatory activity, it can be influenced by current state factors such as recent infections, injuries, body mass index (BMI), or chronic inflammatory conditions ^23^. Evidence also suggests that chronic low grade/sub-acute inflammation over time may be implicated in MDD, which may not be captured by cross-sectional measurements of serum CRP ^8,24^

Recently, a large epigenome wide association study (EWAS) of CRP identified several DNA methylation correlates of low-grade inflammatory activity^25^. From the CpG sites identified in this study, a methylation score for CRP (DNAm CRP) was created ^26^; the score has since been generated in participants of Generation Scotland (n=7,028)^28^. Since this measure of inflammation may be less prone to the acute effects of cross-sectional measures of serum CRP described above, DNAm CRP may provide a more stable signature of exposure to chronic inflammatory states compared to cross-sectional serum CRP ^29,30^. Indeed, methylation risk scores of other phenotypes including MDD itself have proven discriminatory utility, with comparable/better performance compared to genetic risk scores ^31^.

The current study therefore examines a large community-based sample (n=566-798) to investigate (i) associations between serum CRP and DNAm CRP and MDD symptoms, (ii) associations between both CRP (serum CRP and DNAm CRP) measures and structural imaging phenotypes (TI and diffusion MRI) and (iii) interaction effects between both measures of CRP and MDD diagnosis to determine the differential relationship of these inflammatory markers and imaging associations in depression.

## 2. Methods and Materials

### 2.1 Participants

The participants in this study were recruited as part of the STratifying Resilience and Depression Longitudinally (STRADL) study (2015-2019) which re-contacted participants from the Generation Scotland: Scottish Family Health Study (GS) via post for further assessment of mental health, specifically depression. Full details of the STRADL cohort and GS protocol are published elsewhere ^32-37^ The current sample included a range of 566-798 participants depending on the biomarker/imaging modality investigated (Table 1). As GS is a family-based study we randomly included one participant per family in order to have an unrelated sample.

**Table 1:** Participant Demographics.

| **Variable** | **Unit** | **Descriptor** | **N with T1 Data** | **N with DTI Data** |
| --- | --- | --- | --- | --- |
| **Age** | Years (M, SD) | 59.5 (9.5) | - | - |
| **Sex** | Males (N, %) | 333 (41.7) | - | - |
|  | Females (N, %) | 466 (58.3%) | - | - |
| **BMI** | Mean (SD) | 27.98 (5.39) |  |  |
| **SCID Diagnosis** | All |  | 798 | 766 |
|  | Cases (N,%) | 240 (30) | 240 (30) | 228 (29.8) |
|  | Controls (N, %) | 558 (70) | 558 (70) | 538 (70.2) |
| **Total QIDS Score (whole sample)** | Mean (SD) | 4.63 (3.64) | 798 | 766 |
| **C- reactive Protein** | All | 797 | 797 | 765 |
|  | <4mg/L (N,%) | 649 (81.4) | 649 (81.4) | 622 (81.3) |
|  | ≥4mg/L (N,%) | 148 (18.6) | 148 (18.6) | 143 (18.7) |
| **DNAm CRP Score** | Mean (SD) | -0.012 (0.0008) | 591 | 566 |

\* Age, Sex and BMI descriptive statistics calculated on 798 participants with SCID and QIDS data

Ethical approval for STRADL was formally obtained from the NHS Tayside committee on research (reference 14/SS/0039), and all participants provided their written informed consent.

### 2.2 Clinical Assessment

A full medical history was obtained and updated from previous GS baseline assessment ^34^ and any new diagnoses or medical episodes recorded at the imaging assessment. Health and lifestyle data were also collected, as were physical measurements such as height and weight, from which BMI was derived as a covariate of interest ^32^ Smoking status was collected from GS baseline assessments as were the number of smoking pack years-full details have been reported by Barbu and colleagues ^31^.

Participants in STRADL completed a broad range of tests designed to assess depression incidence/severity. All participants were screened for a lifetime history of MDD. A research version of the Structured Clinical Interview for DSM disorders (SCID) ^38^ was used to assess symptoms of mood disorder. Diagnostic criteria were based on the Diagnostic and Statistical Manual of Mental Disorders (DSM-IV-TR). From this, participants were given a binary score of no history of depression (0) or lifetime episode of depression (1). Using this definition our study had 240 cases of lifetime depression and 558 controls in the T1 sample (Table 1). The Quick Inventory of Depressive Symptomatology (QIDS)^39^, a 16-item questionnaire, was employed in order to assess the severity of current depressive symptoms at the time of assessment among study participants. From this, a total QIDS score was calculated as a measure of current depression severity for analyses purposes. Where there were any significant interactions with QIDS score in analyses, scores were grouped in to <=10 or >10 for visualisation of results. Individuals with a score greater than 10 represent those with moderate to severe depression ^39^.

### 2.3 C-reactive Protein Measurement (CRP)

To obtain serum CRP levels, venepuncture was employed using a butterfly needle kit. Blood was extracted into clot activator gel for serum separation. CRP samples were taken and sent to NHS laboratories (Ninewells Hospital/Aberdeen Royal Infirmary) for analysis. The biochemical assay utilized in CRP analyses possessed a detection threshold of 4 mg/L and CRP levels below the 4mg/L detection threshold were recorded as 0. For analyses purposes, CRP levels were stratified into clinically relevant groups: <4 mg/L (clinically normal) and ≥4 mg/L (clinically elevated)^40^. Blood samples were taken concurrently with imaging analyses and depression measures.

### 2.4 DNAm CRP Score Calculation

Blood samples used to generate the DNAm CRP score were collected at the Generation Scotland baseline appointment (between 2006 and 2011) and DNA methylation was profiled using the Illumina Human-MethylationEPIC BeadChip in two different sets. Pre-processing and quality control steps for both sets of methylation data have previously been fully reported^28,41^.

Full details of the calculation of the DNAm CRP score in GS have been reported previously ^28^. Briefly, methylation beta values were extracted for 6 CpG sites shown to have the strongest evidence of a functional association with serum CRP levels as shown by Lighthart and colleagues (n = 8863 and 4111 of European and African ancestries, respectively) ^25^. One of the CpG sites (cg06126421) in the original study was unavailable in the GS dataset and was therefore not included in the current analysis. The beta values for the six CpG sites associated with serum CRP were then multiplied by their respective regression weights and summed to generate a single score for each STRADL participant ^28^. As all the EWAS regression weights were negative, a higher DNAm CRP score corresponds to a score closer to zero.

### 2.5 MRI Acquisition and Analyses

STRADL participants were scanned at two centres: the Ninewells Hospital in Dundee and at the Aberdeen Royal Infirmary in Aberdeen. Both study centres followed the same protocol including structural sequences ^32^ 3T MRI scans were anonymised at the time of acquisition and only the T1 and diffusion MRI (DTI) data are utilized in this study. Full details of the imaging sequences and parameters are provided in supplementary materials.

T1 structural measures were processed using FreeSurfer version 5.3 ^42^ to quantify the volumes of 14 subcortical structures as well as the volumes, surface area and thickness of 34 cortical regions per hemisphere according to the Desikan-Killany atlas ^43^. Full details of the Freesurfer Quality Control (QC) steps are provided in supplementary materials and have also been reported in full previously^44^. Measures of thickness, surface area and volume were derived for each of the 68 cortical regions. The volumes of 14 subcortical structures – left and right accumbens area, amygdala, caudate nucleus, hippocampus, pallidum, putamen and thalamus – were also extracted from FreeSurfer output. Global measures of cortical volume, surface area and thickness were also derived, as well as 5 summed lobar measures (frontal, parietal, temporal, occipital and cingulate; Table S1). The number of QC edits made per individual were recorded to use as a covariate in statistical analyses.

For DTI data, pre-processing and quality control was performed using standard tools available from FSL (https://fsl.fmrib.ox.ac.uk/fsl/fslwiki). Tract Based Spatial Statistics (TBSS) was carried out according to the ‘The Enhancing NeuroImaging Genetics through Meta-Analysis’ (ENIGMA) Consortium DTI protocol (http://enigma.ini.usc.edu/protocols/dti-protocols/). Region of interest (ROI) extraction analyses were then performed also using ENIGMA protocols to extract fractional anisotropy (FA) and mean diffusivity (MD) measures (http://enigma.ini.usc.edu/protocols/dti-protocols/). White matter tracts were categorised using the Johns-Hopkins University DTI-based white matter atlas^45^. This resulted in 5 unilateral tracts and 19 bilateral tracts, as well as an average measure, for FA and MD. This included ten association fibres, three commissural fibres, eight projection fibres and four thalamic radiations (Table S1).

### 2.6 Statistical Analyses

#### Subcortical/Cortical Measures

For all cortical and subcortical measures, age, sex, imaging batch, number of image edits per individual, hemisphere, assessment centre and standardised intracranial volume (ICV) were set as covariates in mixed-effect linear models. BMI was set as an additional covariate for all CRP analyses and methylation set, smoking status and pack years smoked were included in all DNAm CRP analyses. For unilateral structures and global/lobar measures, a general linear model was applied as above. Hemisphere was controlled for as a repeated measure in all bilateral structural neuroimaging phenotypes, using mixed-effect models.

#### White Matter Integrity

For DTI measures, age, sex, and assessment centre were included as covariates as well as BMI/methylation set/smoking variables where relevant. Global integrity was determined by applying principal component analysis (PCA) on the 24 tracts to extract a latent measure. Scores of the first un-rotated principal component of FA/MD were extracted and set as the dependent variable (proportion of variance explained by the first principal component is provided in Table S2). We then separately examined four subsets of white matter tract for which scores of the first un-rotated principal component were also extracted: (a) association fibres, (b) commissural fibres, (c) projection fibres and (d) thalamic radiations. The tracts included in the four subsets are provided in supplementary materials (Table S1). Finally, we examined each white matter tract individually. Mixed-effect linear models were used for the measures of bilateral white matter tracts correcting for hemisphere as a repeated measure, consistent with above, while general linear models were used for the unilateral midline tracts.

#### Statistical Models

All analyses were conducted using R (version 3.2.3) in a Linux environment. Linear mixed-effects models (function ‘lme’ in R package ‘nlme’) and general linear models (function ‘glm’ in R package ‘stats’) were used to investigate structural brain metrics ^46,47^ False Discovery Rate (FDR) multiple comparison correction was applied to all bilateral/unilateral structures, lobes and white matter tracts, referred to as P_FDR_ in this report, using the ‘p.adjust’ function in R and all betas were standardised. FDR correction was also applied over each sub-analysis. We investigated: (i) associations between serum CRP and DNAm CRP and depression symptoms, (ii) associations between both CRP measures and structural imaging phenotypes (T1 and DTI) and (iii) CRP × MDD interaction effects using both measures of CRP (serum and DNAm), and two measures of depression (case/control and QIDS).

## 3. Results

### 3.1 Demographics

Descriptive statistics of the key variables as well as sample numbers for each analysis in this study are presented in Table 1. Case-control MDD and QIDS associations on the structural imaging metrics across the full sample are also provided in supplementary materials (Table S16-S21).

### 3.2 Associations between serum CRP and DNAm CRP with depression symptoms

Increased serum CRP levels were significantly associated with increased depression symptoms as measured by the total QIDS score (β= 0.073, P_FDR_ = 0.033; Figure 1). For the separate QIDS items, increased serum CRP levels were significantly associated with decreased energy levels (β= 0.101, P_FDR_ = 0.027) and decreased general interest (β= 0.145, P_FDR_ = 6*10^-4^). There were no significant associations between serum CRP and case/control MDD status.

**Figure 1:**
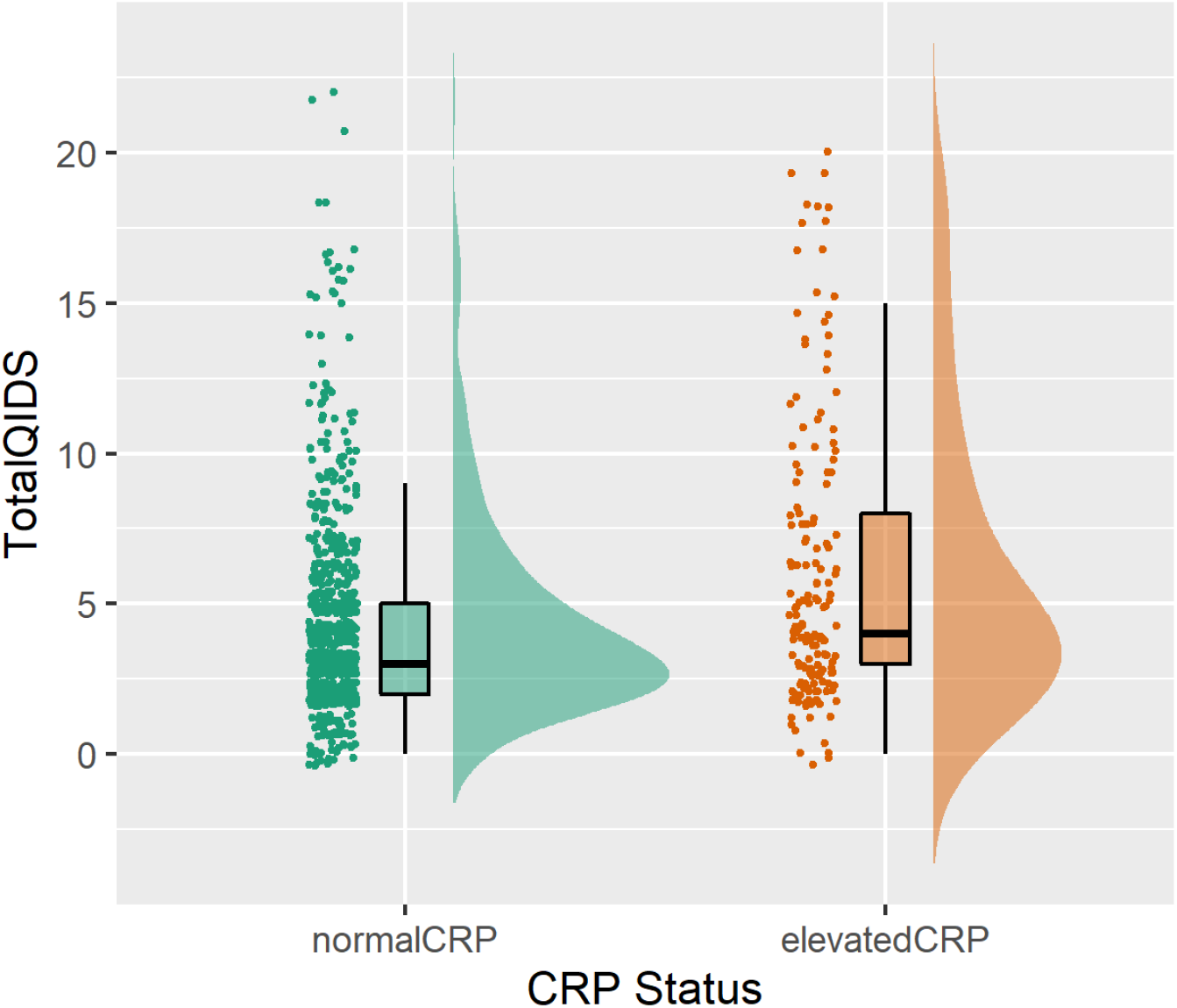
Raincloud plot of associations between serum CRP status and Total QIDS score

There were no significant associations between the DNAm CRP score and any of the measures of depression symptoms or MDD case/control status when controlling for age, sex, methylation set, smoking status, pack years, BMI and assessment centre; however, there were significant associations between DNAm CRP and Total QIDS scores in a minimally adjusted model controlling for age, sex, methylation set and assessment centre (Table S3).

### 3.3 Associations of serum C-reactive Protein and DNAm CRP and structural brain metrics

#### Serum C-reactive Protein

There were no significant associations between serum CRP status and any global measures or lobar measures. For the individual regions, we found elevated CRP status was significantly associated with thinning of the entorhinal cortex (β= −0.095, P_FDR_ = 0.037; Figure 4). There were no significant associations between serum CRP status on any of the DTI measures, globally or regionally.

#### DNAm CRP

For DNAm CRP (n=577), we report a significant association between an increasing DNAm CRP score and smaller global grey matter (β= −0.06, p=0.02) and smaller global cortical volume (β= −0.1, p=0.01; Figure 5). There were no FDR significant associations with any of the regional structural measures indicating DNAm CRP has an effect globally rather than regionally.

With regard to the DTI measures (n=566), increased DNAm CRP scores were significantly associated with differences in white matter microstructure including negative associations with gFA (β= – 0.07, p =0.04), FA in the external capsule (β= −0.14, P_FDR_ = 0.02) and negative associations with FA of the anterior limb of the internal capsule (ALIC; β= −0.12, P_FDR_ = 0.048).

There were also significant associations with MD values for projection fibres (β= 0.098, Pfdr = 0.019) and thalamic radiations (β= 0.079, P_FDR_ = 0.028). We also tested the effects of the DNAm CRP score on individual tracts and found significant effects for tracts summarised in Table 2. The largest effect size found for an individual tract was for the ALIC (β= 0.1, P_FDR_ = 0.042).

**Table 2:** Significant effects of the DNAm CRP score on MD values of DTI Tracts

| White Matter Tract | Mean Diffusivity |  |  |  |  |
| --- | --- | --- | --- | --- | --- |
| | $\beta$ | SE | t value | p value | pFDR |
| Anterior Corona Radiata | 0.068 | 0.042 | 1.608 | 0.108 | 0.134 |
| Anterior Limb Internal Capsule | 0.100 | 0.039 | 2.560 | 0.011 | **0.042** |
| Cingulum (cingulate gyrus) | 0.081 | 0.037 | 2.214 | 0.027 | **0.044** |
| Cingulum (hippocampus) | 0.017 | 0.030 | 0.573 | 0.567 | 0.570 |
| Corona Radiata | 0.091 | 0.040 | 2.305 | 0.022 | **0.042** |
| Corticospinal Tract | 0.091 | 0.039 | 2.344 | 0.019 | **0.042** |
| External Capsule | 0.123 | 0.039 | 3.125 | 0.002 | **0.022** |
| Fornix(Cres)/ Stria Terminalis | 0.079 | 0.032 | 2.457 | 0.014 | **0.042** |
| Internal Capsule | 0.072 | 0.032 | 2.255 | 0.025 | **0.042** |
| Inferior- Fronto-Occipital Fasciculus | 0.084 | 0.037 | 2.257 | 0.024 | **0.042** |
| Posterior Corona Radiata | 0.119 | 0.036 | 3.324 | 0.001 | **0.022** |
| Posterior Limb of Internal Capsule | 0.018 | 0.031 | 0.568 | 0.570 | 0.570 |
| Posterior Thalamic Radiation | 0.077 | 0.037 | 2.058 | 0.040 | 0.057 |
| Retrolenticular part of Internal Capsule | 0.075 | 0.030 | 2.473 | 0.014 | **0.042** |
| Superior Corona Radiata | 0.083 | 0.037 | 2.258 | 0.024 | **0.042** |
| Superior Fronto-Occipital Fasciculus | 0.072 | 0.039 | 1.838 | 0.067 | 0.089 |
| Superior Longitudinal Fasciculus | 0.092 | 0.037 | 2.465 | 0.014 | **0.042** |
| Sagittal Striatum | 0.099 | 0.036 | 2.732 | 0.007 | **0.042** |
| Uncinate Fasciculus | 0.094 | 0.039 | 2.401 | 0.017 | **0.042** |
| Body Corpus Callosum | 0.097 | 0.041 | 2.335 | 0.020 | **0.042** |
| Corpus Callosum | 0.083 | 0.039 | 2.138 | 0.033 | **0.050** |
| Fornix (column and body) | 0.052 | 0.042 | 1.243 | 0.215 | 0.234 |
| Genu Corpus Callosum | 0.063 | 0.040 | 1.571 | 0.117 | 0.134 |
| Splenium of Corpus Callosum | 0.058 | 0.037 | 1.571 | 0.117 | 0.134 |

### 3.4 Interaction effects between peripheral inflammation and depressive symptoms on structural brain metrics

Lastly, we tested interaction effects between both peripheral markers of inflammation and (1) case/control status and (2) total QIDS score on all of the structural brain metrics investigated in this study.

#### Inflammation*MDD Case-Control Interaction Effects

For serum CRP status, there were no significant interaction effects with MDD case/control status on any of the brain metrics investigated. We also found no significant interaction effects between the DNAm CRP and MDD case-control status on any of the structural metrics investigated (Table S10-S15).

#### Inflammation* Total QIDS Interaction Effects

There were no significant interactions between serum CRP status and total QIDS on any cortical or subcortical structures. There was a significant interaction effect between CRP status and total QIDS score for the thickness of the occipital lobe such that those with an increased serum CRP level and increased QIDS score had increased thickness (β= 0.022, P_FDR_ = 0.018; Figure 3). There was however no significant interaction between the DNAm CRP score and total QIDS score on any of the cortical or subcortical structural measures. There was one significant interaction effect with DNAm CRP and total QIDS score on global total grey matter (β= 0.02, P_FDR_ = 0.04) such that individuals with increased QIDS scores and an increased DNAm CRP score have smaller global grey matter volumes (Figure 2).

**Figure 2:**
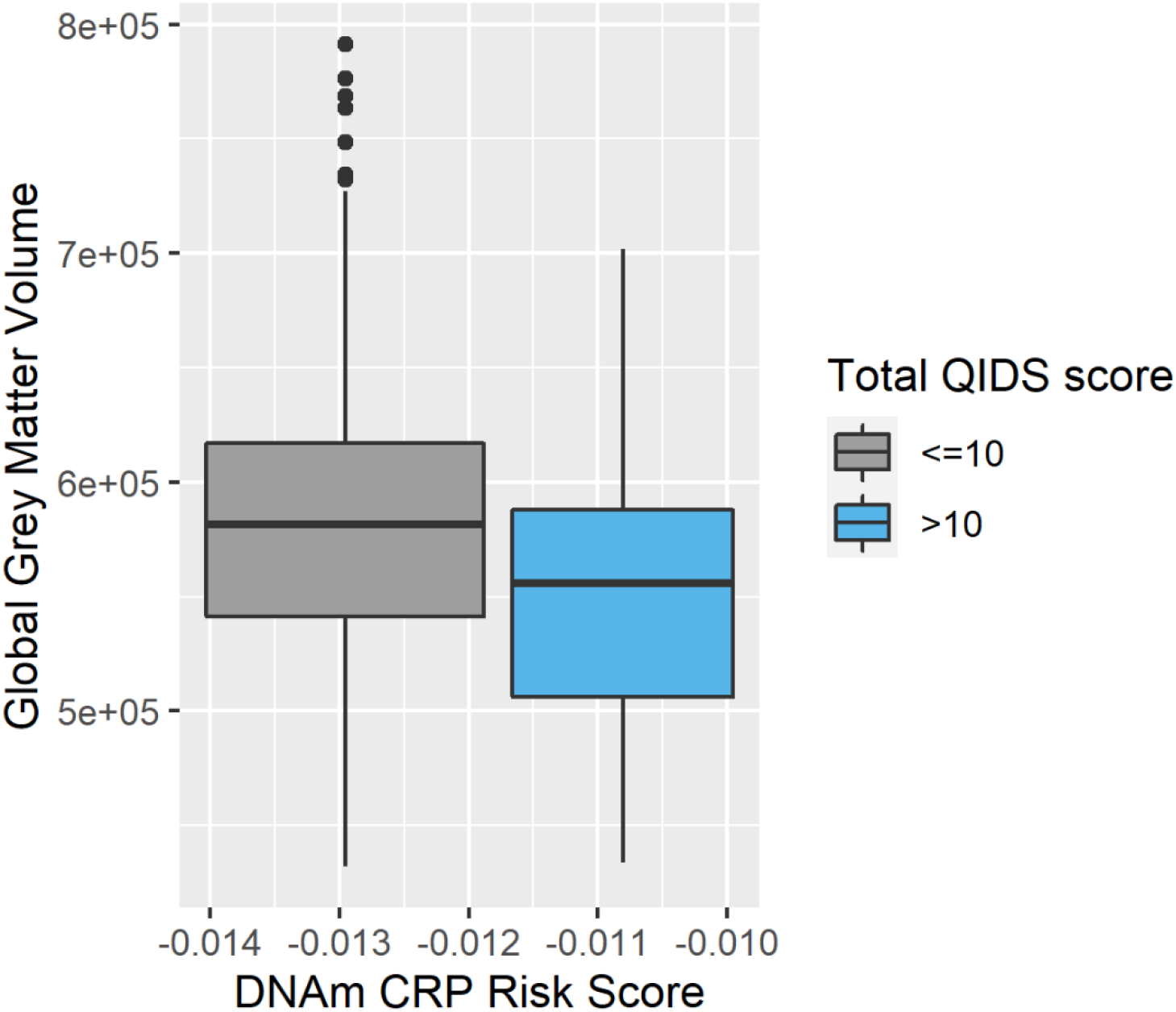
Association between DNAm CRP score, Total QIDS score and Global Grey Matter Volumes

**Figure 3:**
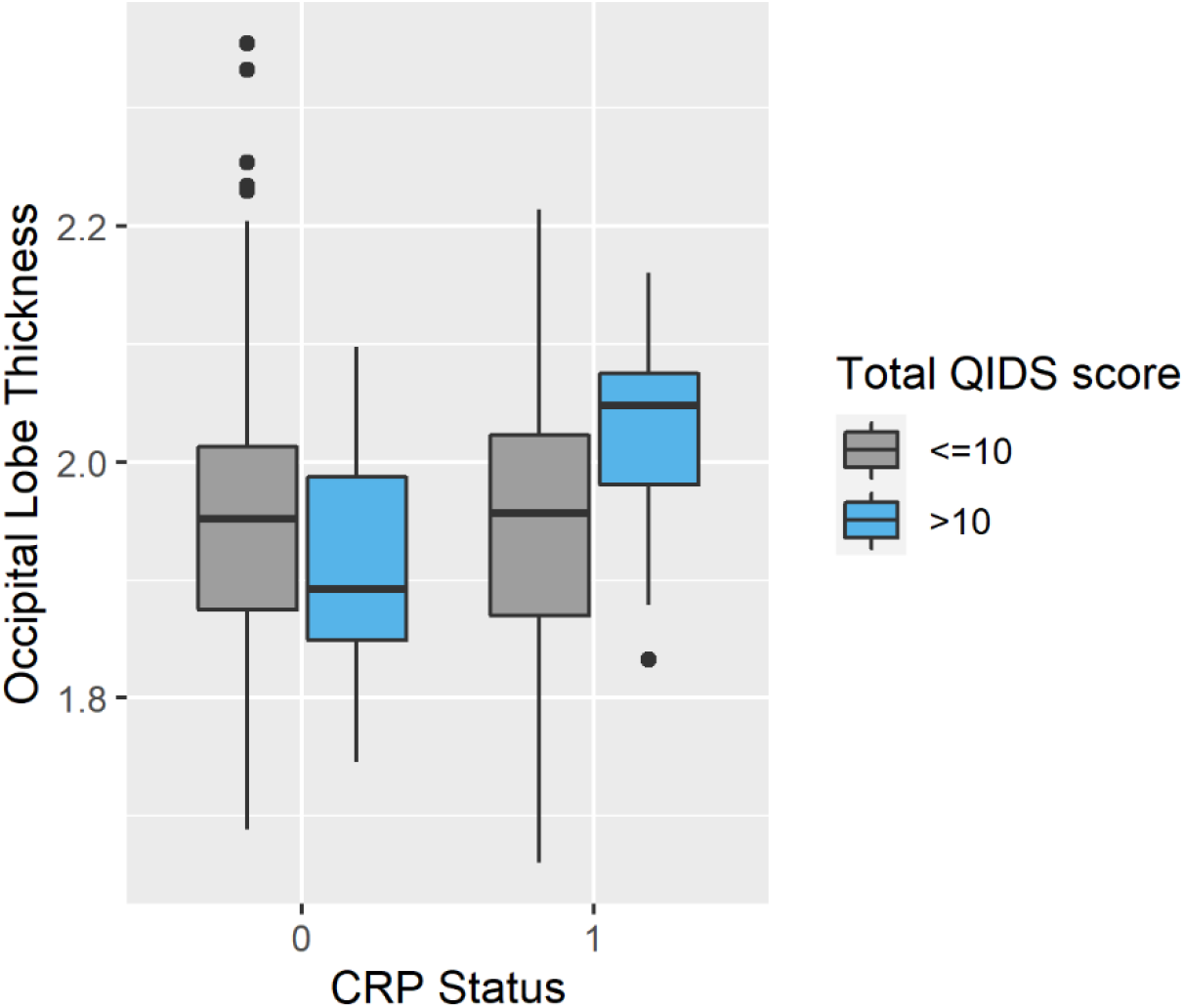
Association between serum CRP status, QIDS Score and Occipital Lobe Thickness. **Fig 3-** CRP Status is binarized as 0= serum CRP <4mg/L and 1= serum CRP ≥4mg/L. QIDS scores have been binarized as 0= QIDS score ≤10, 1= QIDS score >10.

**Figure 4:**
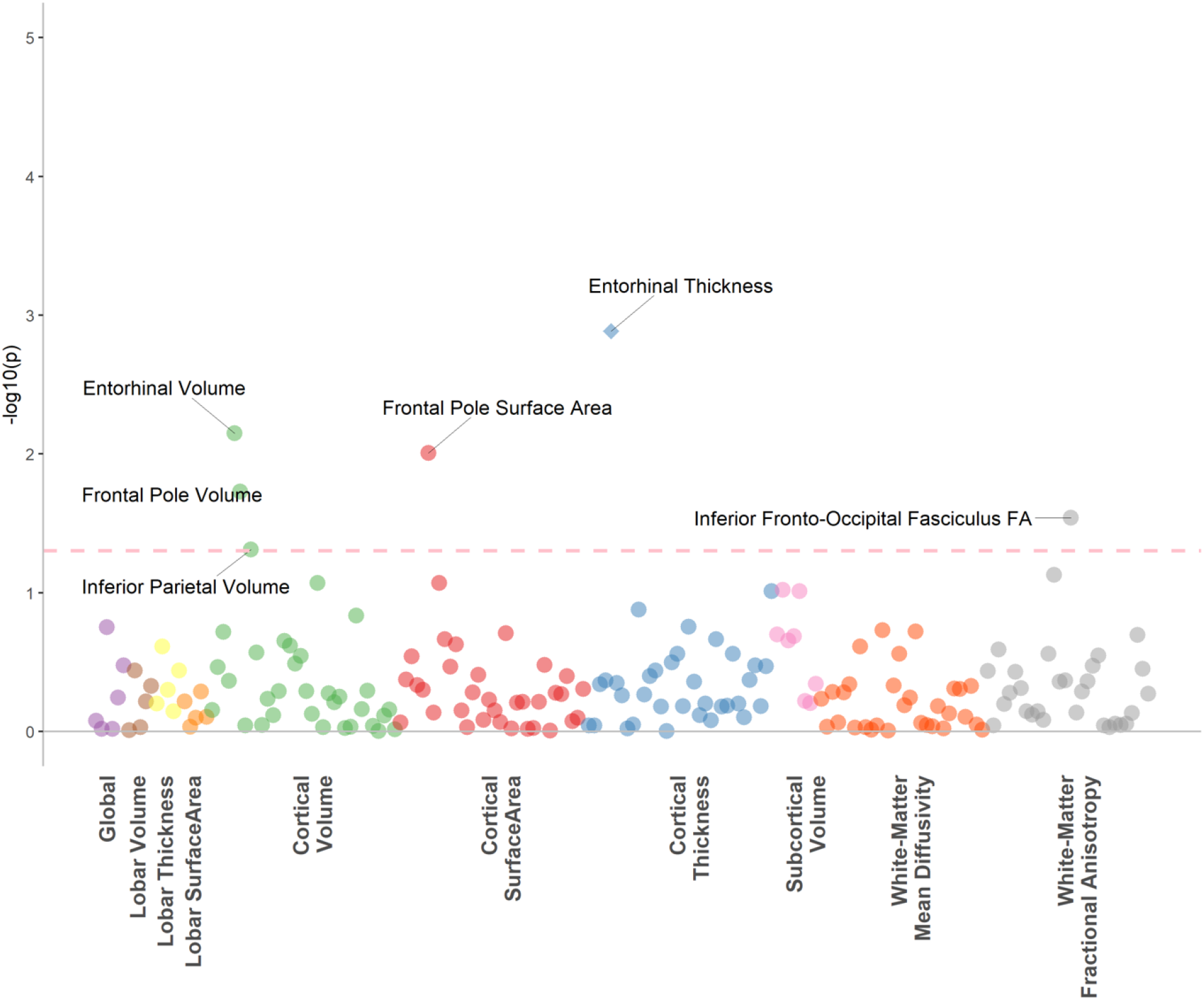
Associations between serum CRP and structural brain phenotypes **Figure 4:** The dotted line indicates the p value threshold 0.05. Each dot represents one structural brain phenotype. Each colour represents one imaging modality. The diamonds represent phenotypes that are also significant after FDR correction.

**Figure 5:**
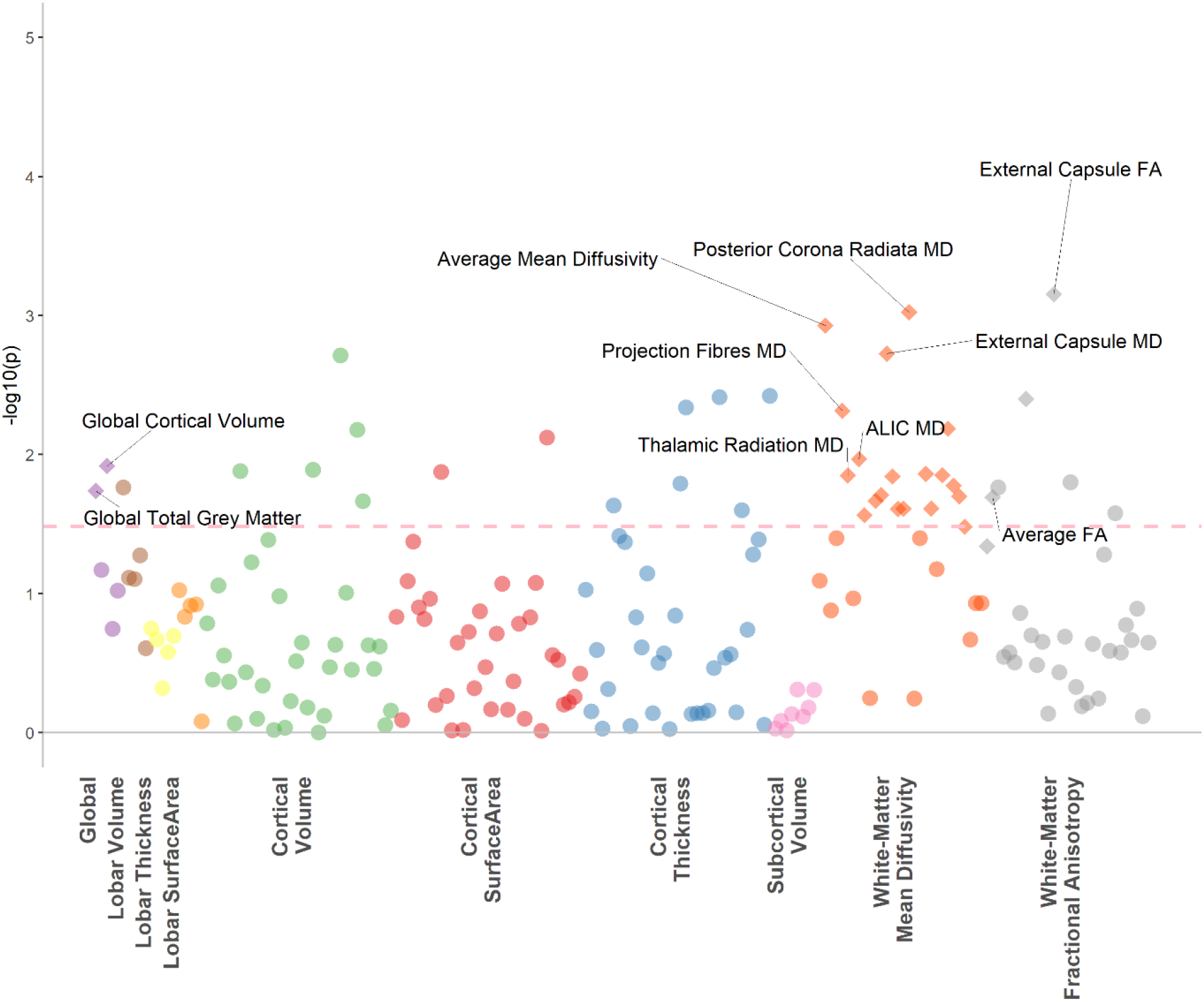
Associations between DNAm CRP score and structural brain phenotypes **Figure 5:** The dotted line indicates the p value threshold 0.05. Each dot represents one structural brain phenotype. Each colour represents one imaging modality. The diamonds represent phenotypes that are also significant after FDR correction.

## 4. Discussion

This study investigated the association between two peripheral markers of CRP and their associations with structural neuroimaging measures and their interaction effects with MDD diagnosis and symptoms. We also investigated the effects of (1) MDD case/control status and (2) total QIDS score on structural imaging measures to characterise the effects of depression on structural imaging measures in our sample before looking at CRP associations.

Across the whole sample, we report that higher serum C-reactive protein was associated with a thinner entorhinal cortex. This ROI has been associated with CRP in previous research. A study by Bettcher and colleagues found CRP was associated with smaller left medial temporal lobe volumes which included the entorhinal cortex, and found that those with detectable levels of CRP demonstrated poorer performance in cognitive tasks ^48^. This area has also been associated with the development of somatic symptoms in previous research^49^ and the medial temporal lobe is thought to be involved in the emotional component of somatic complaints^50^. Previous research on the effects of inflammation on the entorhinal cortex are primarily pre-clinical or in the context of Alzheimer’s disease (AD); however, chronic neuroinflammation is associated with neuronal loss and BBB leakage in this region and the entorhinal cortex may represent an area that is vulnerable to the effects of inflammation ^51,52^

Increased serum CRP levels were significantly associated with increased depression symptoms as measured by the total QIDS score and were significantly associated with two somatic symptoms-general interest and energy levels. These findings are consistent with the existing literature: other studies of inflammation and depression symptoms have found that inflammatory markers are associated with somatic/neurovegetative symptoms of depression including fatigue, impaired sleep and activity rather than psychological symptoms ^53-56^. This suggests that higher cross-sectional serum CRP levels are associated with the somatic symptoms of depression, although we did not find evidence that this was related to white matter integrity or any regional brain structure. Therefore, cross-sectional serum CRP seems to be associated with more dynamic measures such as MDD symptoms rather than long-term structural brain change.

To our knowledge, there are no studies on the relationship between peripheral blood DNA methylation CRP scores and brain structure in the context of depression. We found that DNAm CRP scores had a greater number of associations with imaging traits, with larger effect sizes compared to serum CRP. DNAm CRP scores were associated with several structural neuroimaging measures including reductions in global grey matter and global cortical volume. We also found that increased DNAm CRP scores were associated with white matter changes including reductions in FA in the external capsule and ALIC and increases in MD for projection fibres and thalamic radiations. A study by Freytag and colleagues identified a peripheral blood epigenetic signature associated with cortical thickness and found that the genomic location of the contributing methylation sites mapped to genes involved in the immune system and inflammatory response ^57^ This suggests a possible relationship between methylation markers of the immune system and neuroimaging traits and highlights the utility of methylomic profiles for investigating brain phenotypes.

The current findings contribute to the “inflammaging” theory of ageing, whereby chronic low-grade inflammation accelerates age-related neurodegenerative processes including reductions in cortical volume and white matter integrity ^58 59^. We found that the DNAm score was associated with reductions in both cortical volume and white matter integrity. The reductions in FA associated with DNAm CRP are similar to findings from studies investigating the effect of high-sensitivity serum CRP and white matter integrity ^60^. Furthermore, a longitudinal study of serum CRP over 21 years found that individuals with increasing inflammation after midlife also tended to have greater white matter microstructural abnormalities ^61^. We found that the DNAm CRP score did not have any interaction effects with case/control MDD status or depression symptoms with any of the regional, lobar or DTI neuroimaging measures. However, there was an interaction effect with the DNAm CRP score on depressive symptoms and global grey matter such that individuals with both increased DNAm CRP and depression symptoms had significantly smaller global grey matter volumes. Several potential mechanisms have linked immunobiological changes to affective neurobiology including accelerated biological ageing, such as immune cell senescence, which has well-documented effects on both the epigenome and transcriptome. MDD is associated with immune cell senescence and a study by Diniz et al found that a senescence associated secretory phenotype was associated with MDD severity^62^. Future research could examine the relationships that link epigenome, transcriptome and inflammatory proteins with neuronal damage and subsequent changes in structural imaging phenotypes.

This study has a number of strengths, in particular, the STRADL study is a large community-based sample with in-depth phenotypic assessment and neuroimaging data (n=566-798 in the current study). Our study also benefits from the inclusion of methylation data which provided a long-term signature of chronic/low-level inflammation, overcoming standard issues with single timepoint CRP measurement which fluctuates in response to numerous factors and is prone to measurement error. We have also utilized an unbiased data-driven approach by not selecting neuroimaging regions of interest prior to analyses. This allowed an assessment of the effects on inflammation and depression on a wide range of structural neuroimaging measures.

Limitations of this study included a small number of current MDD cases in comparison to the number of controls. As this study is a community-based sample, this indicates that the participants with MDD are relatively well in comparison to MDD cases who are hospitalised. Therefore, it is possible that we are not capturing the effects of the most severe forms of MDD. However, community-based sampling results may be more generalizable to the population than those obtained in a clinic. Furthermore, although CRP is widely used clinically to determine peripheral inflammation, it is ultimately a proxy for inflammatory activity^28^ and the detection threshold for CRP in this study was 4mg/L; values lower than this were recorded as 0. This could lead to missing biologically relevant interactions at lower, but still clinically relevant, levels of CRP as CRP is known to have consequences on health and lifestyle factors at smaller concentrations. For example, the American Heart Association considers CRP concentrations between 1-3 mg/L to be associated with moderate risk for cardiovascular disease ^40^. Therefore, we note that our study is potentially not capturing the low-level inflammation that may be associated with structural brain changes. However, the insights gained by investigating more acute levels of CRP are still valuable to further our understanding of the effect of inflammatory states on neurobiological structure and function. Lastly, although our study has found associations between peripheral inflammatory markers and structural brain alterations, the study does not provide evidence for causal mechanistic pathways.

In conclusion, this study highlights the need to capture inflammatory activity longitudinally as chronic elevations of inflammation are likely to be involved in structural brain changes and the pathophysiology of psychiatric outcomes ^58,63,64^ We found that serum CRP was associated with depression symptoms, in particular somatic symptoms, and reduction of entorhinal cortex thickness. This study used a methylomic signature of C-Reactive Protein to capture chronic signatures of low-level inflammation and found an association with several structural neuroimaging measures, in particular, differences in white matter integrity. This methylation signature also had significant interaction effects with depression symptoms on global grey matter – providing further support for the role of chronic inflammation in the pathophysiology of major depressive disorder. This study also highlights the utility of using both serological and methylation markers in a multi-level approach to study brain imaging and psychiatric phenotypes.

## Data Availability

Access to and use of GS and STRADL data must be approved by the GS Access Committee under the terms of consent. Full details of the application process can be found at www.generationscotland.org

## Acknowledgements

The authors thank all of the STRADL and Generation Scotland participants for their time and effort taking part in this study. We would also like to thank all of the research assistants, clinicians and technicians for their help in the collecting this data.

## References

1. Whiteford, H. A. et al. Global burden of disease attributable to mental and substance use disorders: Findings from the Global Burden of Disease Study 2010. Lancet (2013) doi:10.1016/S0140-6736(13)61611-6.

2. Sinyor, M., Rezmovitz, J. & Zaretsky, A. Screen all for depression. BMJ (Online) (2016) doi:10.1136/bmj.i1617.

3. Marcus, M., Yasamy, M. T., van Ommeren, M. & Chisholm, D. Depression, a global public health concern. WHO Department of Mental Health and Substance Abuse (2012).

4. Bhattacharya, A., Derecki, N. C., Lovenberg, T. W. & Drevets, W. C. Role of neuro-immunological factors in the pathophysiology of mood disorders. Psychopharmacology (2016) doi:10.1007/s00213-016-4214-0.

5. Dantzer, R., O’Connor, J. C., Freund, G. G., Johnson, R. W. & Kelley, K. W. From inflammation to sickness and depression: When the immune system subjugates the brain. Nature Reviews Neuroscience (2008) doi:10.1038/nrn2297.

6. Strawbridge, R. et al. Inflammation and clinical response to treatment in depression: A meta-analysis. European Neuropsychopharmacology (2015) doi:10.1016/j.euroneuro.2015.06.007.

7. Raison, C. L. et al. A randomized controlled trial of the tumor necrosis factor antagonist infliximab for treatment-resistant depression: The role of baseline inflammatory biomarkers. Arch. Gen. Psychiatry (2013) doi:10.1001/2013.jamapsychiatry.4.

8. Haapakoski, R., Mathieu, J., Ebmeier, K. P., Alenius, H. & Kivimäki, M. Cumulative meta-analysis of interleukins 6 and 1β, tumour necrosis factor α and C-reactive protein in patients with major depressive disorder. Brain. Behav. Immun. (2015) doi:10.1016/j.bbi.2015.06.001.

9. Lindqvist, D. et al. Interleukin-6 Is Elevated in the Cerebrospinal Fluid of Suicide Attempters and Related to Symptom Severity. Biol. Psychiatry (2009) doi:10.1016/j.biopsych.2009.01.030.

10. Liu, Y., Ho, R. C. M. & Mak, A. Interleukin (IL)-6, tumour necrosis factor alpha (TNF-α) and soluble interleukin-2 receptors (sIL-2R) are elevated in patients with major depressive disorder: A meta-analysis and meta-regression. Journal of Affective Disorders (2012) doi:10.1016/j.jad.2011.08.003.

11. Pandey, G. N. et al. Proinflammatory cytokines in the prefrontal cortex of teenage suicide victims. J. Psychiatr. Res. (2012) doi:10.1016/j.jpsychires.2011.08.006.

12. Dowlati, Y. et al. A Meta-Analysis of Cytokines in Major Depression. Biol. Psychiatry (2010) doi:10.1016/j.biopsych.2009.09.033.

13. Kappelmann, N., Lewis, G., Dantzer, R., Jones, P. B. & Khandaker, G. M. Antidepressant activity of anti-cytokine treatment: A systematic review and metaanalysis of clinical trials of chronic inflammatory conditions. Mol. Psychiatry (2018) doi:10.1038/mp.2016.167.

14. Lowe, G. D. O. Circulating inflammatory markers and risks of cardiovascular and non-cardiovascular disease. in Journal of Thrombosis and Haemostasis (2005). doi:10.1111/j.1538-7836.2005.01416.x.

15. Osimo, E. F., Baxter, L. J., Lewis, G., Jones, P. B. & Khandaker, G. M. Prevalence of low-grade inflammation in depression: A systematic review and meta-Analysis of CRP levels. Psychological Medicine (2019) doi:10.1017/S0033291719001454.

16. Valkanova, V., Ebmeier, K. P. & Allan, C. L. CRP, IL-6 and depression: A systematic review and meta-analysis of longitudinal studies. Journal of Affective Disorders (2013) doi:10.1016/j.jad.2013.06.004.

17. Au, B., Smith, K. J., Gariépy, G. & Schmitz, N. The longitudinal associations between C-reactive protein and depressive symptoms: Evidence from the English Longitudinal Study of Ageing (ELSA). Int. J. Geriatr. Psychiatry (2015) doi:10.1002/gps.4250.

18. Chamberlain, S. R. et al. Treatment-resistant depression and peripheral C-reactive protein. Br. J. Psychiatry (2019) doi:10.1192/bjp.2018.66.

19. Quan, N. & Banks, W. A. Brain-immune communication pathways. Brain, Behavior, and Immunity (2007) doi:10.1016/j.bbi.2007.05.005.

20. Elwood, E., Lim, Z., Naveed, H. & Galea, I. The effect of systemic inflammation on human brain barrier function. Brain. Behav. Immun. (2017) doi:10.1016/j.bbi.2016.10.020.

21. Kuhlmann, C. R. W. et al. Mechanisms of C-reactive protein-induced blood–brain barrier disruption. Stroke (2009) doi:10.1161/STROKEAHA.108.535930.

22. Opel, N. et al. Large-scale evidence for an association between low-grade peripheral inflammation and brain structural alterations in major depression in the bidirect study. J. Psychiatry Neurosci. (2019) doi:10.1503/jpn.180208.

23. Kathiresan, S. et al. Contribution of clinical correlates and 13 C-reactive protein gene polymorphisms to interindividual variability in serum C-reactive protein level. Circulation (2006) doi:10.1161/CIRCULATIONAHA.105.591271.

24. Goldsmith, D. R., Rapaport, M. H. & Miller, B. J. A meta-analysis of blood cytokine network alterations in psychiatric patients: Comparisons between schizophrenia, bipolar disorder and depression. in Molecular Psychiatry (2016). doi:10.1038/mp.2016.3.

25. Ligthart, S. et al. DNA methylation signatures of chronic low-grade inflammation are associated with complex diseases. Genome Biol. (2016) doi:10.1186/s13059-016-1119-5.

26. Barker, E. D. et al. Inflammation-related epigenetic risk and child and adolescent mental health: A prospective study from pregnancy to middle adolescence. Dev. Psychopathol. (2018) doi:10.1017/S0954579418000330.

27. Stevenson, A. J. et al. Characterisation of an inflammation-related epigenetic score and its association with cognitive ability. bioRxiv (2019) doi:10.1101/802009.

28. Stevenson, A. J. et al. Characterisation of an inflammation-related epigenetic score and its association with cognitive ability. Clin. Epigenetics 12, 113 (2020).

29. Byun, H. M. et al. Temporal stability of epigenetic markers: Sequence characteristics and predictors of short-term DNA methylation variations. PLoS One (2012) doi:10.1371/journal.pone.0039220.

30. Talens, R. P. et al. Variation, patterns, and temporal stability of DNA methylation: considerations for epigenetic epidemiology. FASEB J. (2010) doi:10.1096/fj.09-150490.

31. Barbu, M. C. et al. Epigenetic prediction of major depressive disorder. Mol. Psychiatry (2020) doi:10.1038/s41380-020-0808-3.

32. Habota, T. et al. Cohort profile for the STratifying Resilience and Depression Longitudinally (STRADL) study: A depression-focused investigation of Generation Scotland, using detailed clinical, cognitive, and neuroimaging assessments. Wellcome Open Res. (2019) doi:10.12688/wellcomeopenres.15538.1.

33. Navrady, L. B. et al. Cohort profile: Stratifying Resilience and Depression Longitudinally (STRADL): A questionnaire follow-up of Generation Scotland: Scottish Family Health Study (GS: SFHS). Int. J. Epidemiol. (2018) doi:10.1093/ije/dyx115.

34. Smith, B. H. et al. Cohort profile: Generation scotland: Scottish family health study (GS: SFHS). The study, its participants and their potential for genetic research on health and illness. Int. J. Epidemiol. (2013) doi:10.1093/ije/dys084.

35. Rupprechter, S. et al. Blunted medial prefrontal cortico-limbic reward-related effective connectivity and depression. Brain (2020) doi:10.1093/brain/awaa106.

36. Romaniuk, L. et al. The Neurobiology of Personal Control During Reward Learning and Its Relationship to Mood. Biol. Psychiatry Cogn. Neurosci. Neuroimaging (2019) doi:10.1016/j.bpsc.2018.09.015.

37. Stolicyn, A. et al. Automated classification of depression from structural brain measures across two independent community-based cohorts. Hum. Brain Mapp. n/a, (2020).

38. First, M. B., Spitzer, R. L., Gibbon, M. & Williams, J. B. W. Structured Clinical Interview for DSM-IV-TR Axis I Disorders, Patient Edition (SCID-I/P, 11/2002 revision). for DSMIV (2002). doi:M.

39. John Rush, A. et al. The inventory for depressive symptomatology (IDS): Preliminary findings. Psychiatry Res. (1986) doi:10.1016/0165-1781(86)90060-0.

40. Ridker, P. M. Clinical application of C-reactive protein for cardiovascular disease detection and prevention. Circulation (2003) doi:10.1161/01.CIR.0000053730.47739.3C.

41. Madden, R. A. et al. Birth weight associations with psychiatric and physical health, cognitive function, and DNA methylation differences in an adult population. *bioRxiv* 664045 (2020) doi:10.1101/664045.

42. Dale, A. et al. FreeSurfer Manual. Neuroimage (2002).

43. Desikan, R. S. et al. An automated labeling system for subdividing the human cerebral cortex on MRI scans into gyral based regions of interest. Neuroimage (2006) doi:10.1016/j.neuroimage.2006.01.021.

44. Neilson, E. et al. Impact of Polygenic Risk for Schizophrenia on Cortical Structure in UK Biobank. Biol. Psychiatry (2019) doi:10.1016/j.biopsych.2019.04.013.

45. Mori, S., Van Zijl, P. & Tamminga, C. A. Human white matter atlas. Am. J. Psychiatry (2007) doi:10.1176/ajp.2007.164.7.1005.

46. Shen, X. et al. Subcortical volume and white matter integrity abnormalities in major depressive disorder: Findings from UK Biobank imaging data. Sci. Rep. (2017) doi:10.1038/s41598-017-05507-6.

47. Shen, X. et al. A phenome-wide association and Mendelian Randomisation study of polygenic risk for depression in UK Biobank. Nat. Commun. 11, 2301 (2020).

48. Bettcher, B. M. et al. C-reactive protein is related to memory and medial temporal brain volume in older adults. Brain. Behav. Immun. (2012) doi:10.1016/j.bbi.2011.07.240.

49. Wei, D. et al. Regional gray matter volume and anxiety-related traits interact to predict somatic complaints in a non-clinical sample. Soc. Cogn. Affect. Neurosci. (2015) doi:10.1093/scan/nsu033.

50. Phelps, E. A. Emotion and Cognition: Insights from Studies of the Human Amygdala. Annu. Rev. Psychol. (2006) doi:10.1146/annurev.psych.56.091103.070234.

51. Hauss-Wegrzyniak, B., Lynch, M. A., Vraniak, P. D. & Wenk, G. L. Chronic brain inflammation results in cell loss in the entorhinal cortex and impaired LTP in perforant path-granule cell synapses. Exp. Neurol. (2002) doi:10.1006/exnr.2002.7966.

52. Montagne, A. et al. APOE4 leads to blood-brain barrier dysfunction predicting cognitive decline. Nature (2020) doi:10.1038/s41586-020-2247-3.

53. Chu, A. L. et al. Longitudinal association between inflammatory markers and specific symptoms of depression in a prospective birth cohort. Brain. Behav. Immun. (2019) doi:10.1016/j.bbi.2018.11.007.

54. Jokela, M., Virtanen, M., Batty, G. D. & Kivimäki, M. Inflammation and Specific Symptoms of Depression. JAMA Psychiatry (2016) doi:10.1001/jamapsychiatry.2015.1977.

55. Köhler-Forsberg, O. et al. Association between C-reactive protein (CRP) with depression symptom severity and specific depressive symptoms in major depression. Brain. Behav. Immun. (2017) doi:10.1016/j.bbi.2017.02.020.

56. Duivis, H. E., Vogelzangs, N., Kupper, N., De Jonge, P. & Penninx, B. W. J. H. Differential association of somatic and cognitive symptoms of depression and anxiety with inflammation: Findings from the netherlands study of depression and anxiety (NESDA). Psychoneuroendocrinology (2013) doi:10.1016/j.psyneuen.2013.01.002.

57. Freytag, V. et al. A peripheral epigenetic signature of immune system genes is linked to neocortical thickness and memory. Nat. Commun. (2017) doi:10.1038/ncomms15193.

58. Franceschi, C. & Campisi, J. Chronic inflammation (Inflammaging) and its potential contribution to age-associated diseases. Journals of Gerontology – Series A Biological Sciences and Medical Sciences (2014) doi:10.1093/gerona/glu057.

59. Zhao, L. et al. Age-Related Differences in Brain Morphology and the Modifiers in Middle-Aged and Older Adults. Cereb. Cortex (2019) doi:10.1093/cercor/bhy300.

60. Wersching, H. et al. Serum C-reactive protein is linked to cerebral microstructural integrity and cognitive function. Neurology (2010) doi:10.1212/WNL.0b013e3181d7b45b.

61. Walker, K. A. et al. The association of mid-to late-life systemic inflammation with white matter structure in older adults: The Atherosclerosis Risk in Communities Study. Neurobiol. Aging (2018) doi:10.1016/j.neurobiolaging.2018.03.031.

62. Diniz, B. S., Reynolds, C. F., Sibille, E., Bot, M. & Penninx, B. W. J. H. Major depression and enhanced molecular senescence abnormalities in young and middle-aged adults. Transl. Psychiatry (2019) doi:10.1038/s41398-019-0541-3.

63. Cunningham, C. & Hennessy, E. Co-morbidity and systemic inflammation as drivers of cognitive decline: New experimental models adopting a broader paradigm in dementia research. Alzheimer’s Research and Therapy (2015) doi:10.1186/s13195-015-0117-2.

64. Miller, A. H. & Raison, C. L. The role of inflammation in depression: From evolutionary imperative to modern treatment target. Nature Reviews Immunology (2016) doi:10.1038/nri.2015.5.

